# Spreading of COVID-19 in Italy as the spreading of a wave packet

**DOI:** 10.1101/2020.08.03.20167197

**Authors:** A. Feoli, A. L. Iannella, E. Benedetto

## Abstract

We find that the spreading of the COVID-19 pandemic in Italy can be described as the propagation of a wave packet in a dispersive medium where the effect of Lockdown is simulated by the dispersion relation of the medium. We start expanding a previous statistical analysis based on the official data provided by the Italian civil protection during 100 days, from March 2nd to June 9th. As the total number of people infected with the virus is uncertain, we have considered the trend of ICU patients and the sum of hospitalized patients and the deceased. Both the corresponding curves are well approximated by the same function depending on four free parameters. The model allows to predict the short term behavior of the pandemic and to estimate the benefits due to lockdown measures.

## 1 Introduction

Unfortunately, in late January of 2020 the first two cases of COVID-19 in Italy appeared. From that moment the virus began to spread with a rapid growth of ICU (Intensive Care Unit) patients. On February 21st there was the first case of death and, at the beginning of March, all Italian regions had been infected. For this reason, the Italian Government had to adopt a series of restrictive measures. On March 5th all Schools and Universities in Italy closed their buildings to the students. Indeed, from March 8th, the population was forced to respect the quarantine and therefore it was possible to leave the house only in cases of extreme necessity. The lockdown initially involved only some “red zones” of northern Italy. Subsequently, the spread of the virus caused the extension of the restrictive measures to the whole Nation. In a previous paper [1], we have analyzed the official data provided by the Italian Civil Protection, starting from March 2nd to May 1st [2] (note that the number of total deaths registered in the table of March 26th were corrected from 8165 to 8215 as suggested by Italian Civil Protection), now we want to analyze the same data until June 9th. It should be kept in mind that lockdown lasted until May 4th, when the so-called “phase 2” began, but a lot of less restrictive measures have started from May 18th. Hence we can consider that the effects of lockdown finish two weeks later, at the beginning of June.

Simultaneously with the spread of the virus, it has become important to analyze mathematically the data not only in Italy but also in other countries [3-8]. It is obvious that the total number of people infected with COVID-19 is impossible to be known exactly. Indeed, there are many people infected but asymptomatic and, if they have not been subjected to the virus test, they are not counted in the official data. This can also be understood by analyzing the percentage of deaths compared to the infected. We currently have, in Italy, a mortality percentage of over 14% and this is caused by the high number of hidden infected. If we knew the real high number of people who contracted the virus, obviously the mortality rate would decrease considerably. Therefore, we have focused on the data which are less uncertain, analyzing the time evolution of the total of ICU patients in Italy and the sum of hospitalized patients and the deceased (note that ICU patients are included in the number of hospitalized patients). In Sect.2 we propose a suitable function that can fit the experimental data and four figures representing the temporal evolution of the pandemic, in Sect.3 we review the standard spreading of a wave packet, in Sect.4 we use the model to estimate the effect of lockdown on the spreading of COVID-19 and, finally, in Sect.5 we discuss the obtained results.

## 2 A simple model for predictions of pandemic behavior

We have found a four parameters function that describes the intensity of pandemic

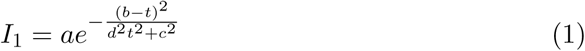

This fitting function can be applied to the curves (already studied in a previous paper [1]) of ICU patients (**Fig. 1**) leading to the values of best fit parameters in **Table 1** and of hospitalized patients plus deaths (**Fig. 2**) with the corresponding results in **Table 2**. We have performed the final fit (**Fig. 1b** and **2b**), but we show also one intermediate fit (**Fig. 1a** and **2a**) to put in evidence how the function (1) can work not only to predict the short term (two or three days) behavior of the pandemic but, in some cases, for long term forecasts as well. For example, in **Figure 1a** the fit made using only the first 45 experimental points (green points in the figure) is able to predict the trend of the curve for all the 55 following days (the prediction done on April 15th works until June 9th), while in **Figure 2a** the fit of the first 50 days predicts well the behavior for the 25 following blue points in the figure (the prediction done on April 20th works until May 15th). We underline also that the curve of “hospitalized patients plus deaths” has a limit in its degrowth imposed by the total number of deaths that, in the plot, plays the role of an horizontal asymptote.

**Table 1.**
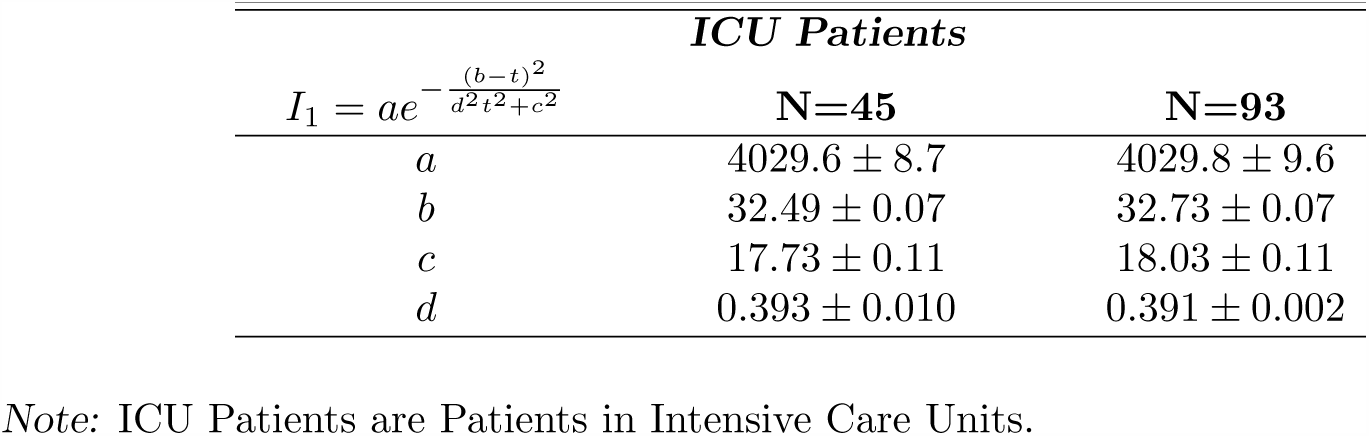
Best fit parameters for the curves in Fig. 1 of ICU Patients of the Epidemic Spreading for 45 and 93 points.

| *ICU Patients* |  |  |
| --- | --- | --- |
| $I_1 = ae^{-\frac{(b-t)^2}{d^2t^2+c^2}}$ | N=45 | N=93 |
| $a$ | $4029.6 \pm 8.7$ | $4029.8 \pm 9.6$ |
| $b$ | $32.49 \pm 0.07$ | $32.73 \pm 0.07$ |
| $c$ | $17.73 \pm 0.11$ | $18.03 \pm 0.11$ |
| $d$ | $0.393 \pm 0.010$ | $0.391 \pm 0.002$ |

Note: ICU Patients are Patients in Intensive Care Units.

**Table 2.** Best fit parameters for the curves in Fig. 2 of H Patients + Deaths of the Epidemic Spreading for 50 and 99 points.

| *H Patients + Deaths* |  |  |
| --- | --- | --- |
| $I_1 = ae^{-\frac{(b-t)^2}{d^2t^2+c^2}}$ | N=50 | N=99 |
| *a* | 51700 ± 119 | 51522 ± 88 |
| *b* | 45.10 ± 0.47 | 45.73 ± 0.11 |
| *c* | 22.25 ± 0.24 | 22.20 ± 0.20 |
| *d* | 0.892 ± 0.036 | 0.970 ± 0.007 |

Note: H Patients are composed by Hospitalized Patients with symptoms and Patients in Intensive Care Units.

**Figure 1a.**
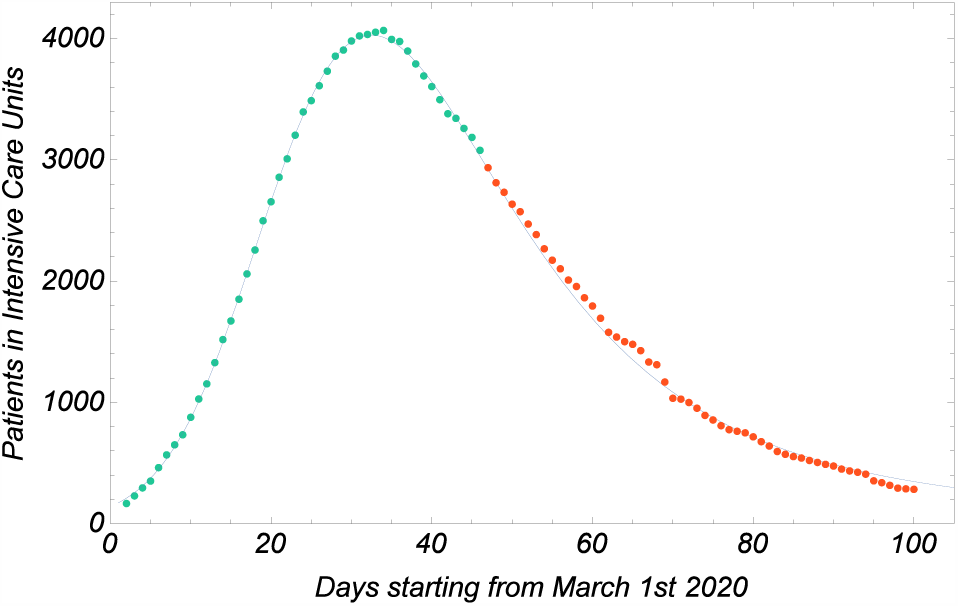
Total number of patients in Intensive Care Units during the examined interval of time. Intermediate fit: the best fit curve (Eq. (1) in the text) is obtained using only the first 45 experimental green points and predicts well the behavior of the remaining 55 points.

**Figure 1b.**
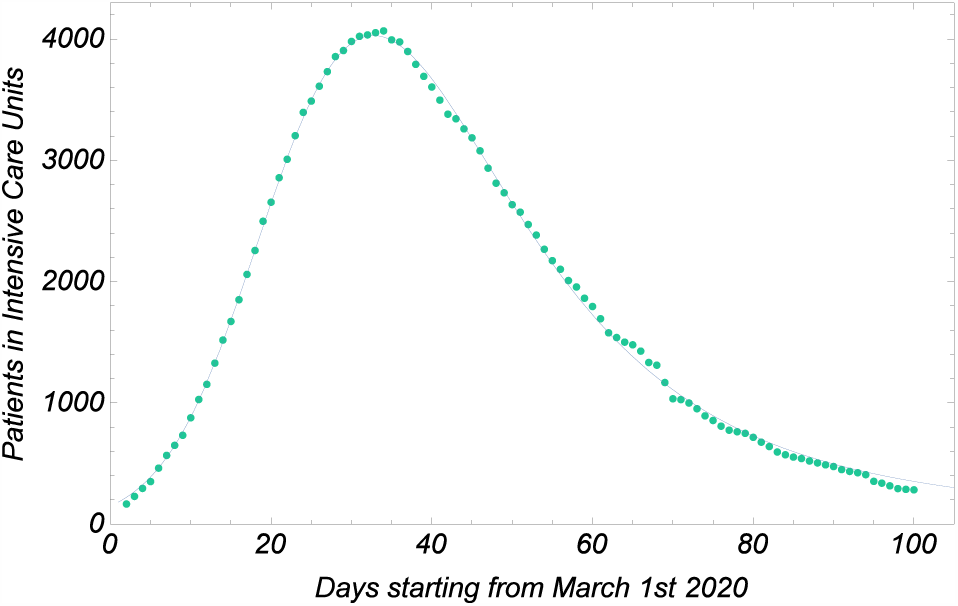
Total number of patients in Intensive Care Units during the examined interval of time. Final fit: the best fit curve (Eq. (1) in the text) is obtained using 93 experimental points

**Figure 2a.**
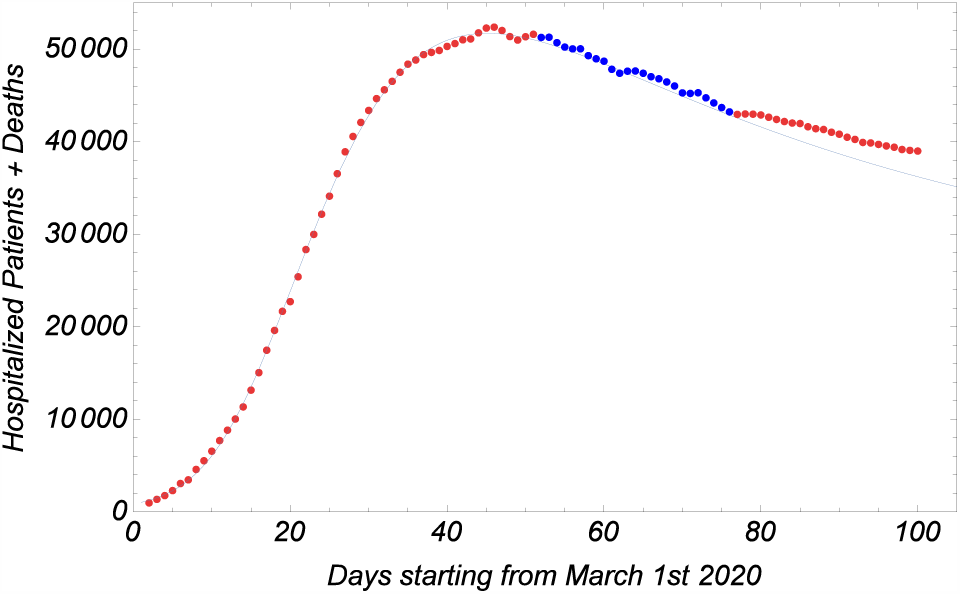
Total number of Hospitalized Patients (Hospitalized Patients with symptoms and Patients in Intensive Care Units) and Deaths. Intermediate fit: the best fit curve (Eq. (1) in the text) is obtained using only the first 50 experimental red points and predicts well the behavior of the following 25 blue points.

**Figure 2b.**
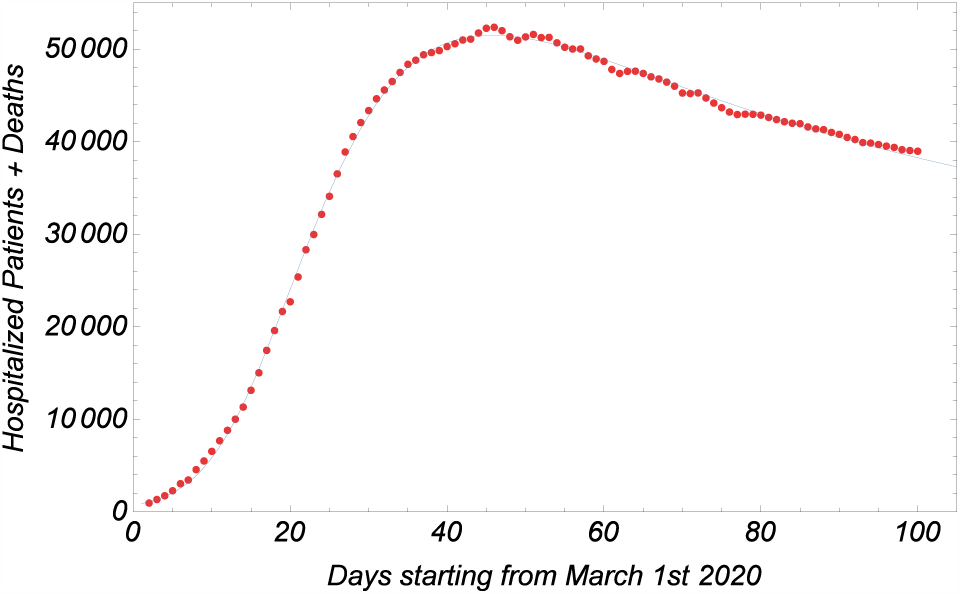
Total number of Hospitalized Patients (Hospitalized Patients with symptoms and Patients in Intensive Care Units) and Deaths. Final fit: the best fit curve (Eq. (1) in the text) is obtained using 99 experimental points.

The width of the gaussian packet increases with time in the typical way of the spreading of a wave packet in a dispersive medium. The same does not occur for the amplitude that does not decrease but remains constant. However, it is worth to examine in details the analogy with the spreading of the wave packet in order to construct a useful model.

## 3 The spreading of a wave packet

In standard textbooks of electromagnetism [9] or quantum mechanics [10], the propagation of a wave packet in a dispersive medium is explained in details. The super-position of a lot of monochromatic waves with wave numbers *k* in a small interval around a central value *k*_0_ is described by the integral

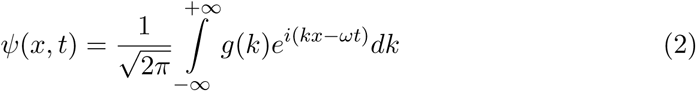

where a gaussian wave packet is characterized by a function

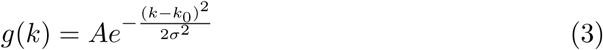

Therefore

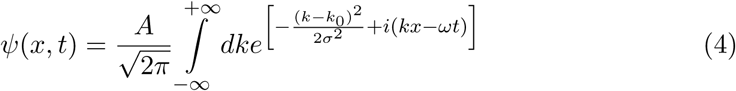

We consider the expansion of the dispersion relation around *k*_0_

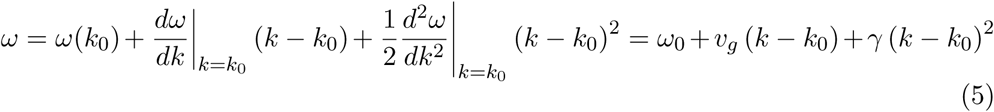

getting

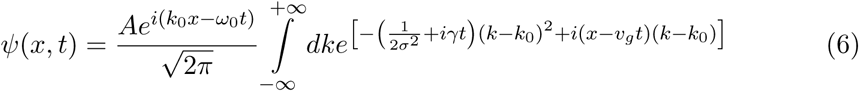

Thanks to the relation

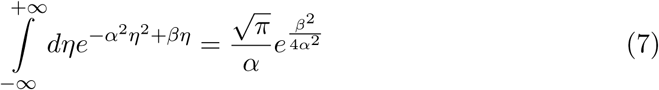

with 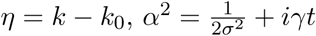 and *β* = *i* (*x* − *v*_*g*_*t*) we can write

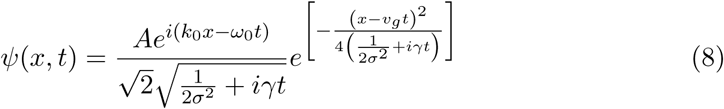

Finally, we obtain

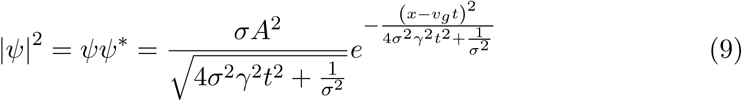

in which the amplitude of the wave packet decreases with time and the width increases with time. These packets are well known in quantum mechanics as solutions of the Schroedinger equation (de Broglie waves). In that framework the equation refers to a particle of mass *m* and the square modulus of the wave function, suitably normalized, is interpreted as the probability density to observe that particle in a point *x* at the instant of time *t* with an experiment. If we consider a beam of *N* identical particles, the wave function allows to predict the subset *n* of particles we will find in a point *x* at an instant *t*.

In order to construct a model to explain the pandemic curves, we can suppose that the spreading of COVID-19 satisfies a mathematical framework similar to the one of Schroedinger wave mechanics. Of course, in this analogy, the lockdown measures play the role of the dispersive medium. The probabilistic approach to the pandemic would allow to predict which fraction *n* of the inhabitants *N* of a nation will become, for example, ICU patients at an instant of time t. This means to consider the intensity of the wave (9) at a constant *x* and to follow its evolution as a function of only one variable: the time. The result is that the function that we must use to fit the experimental data is

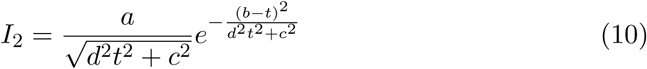

with four free parameters *a* = *A*^2^*σ*/*v*_*g*_, *b* = *x*/*v*_*g*_, *c* = (*σv*_*g*_)^−1^, *d* = 2*γσ*/*v*_*g*_ and, if *a*/*c* must be dimensionless, we have *a,b* and *c* with dimension of time and *d* dimensionless.

## 4 A possible estimation of the effect of Lockdown

We have checked that also the curve in Equation (10) gives a good approximation of the experimental data (**Fig. 3**) and the best fit parameters are in **Table 3**. The very impressive coincidence of the curve predicted by the model with the experimental points was not a foregone conclusion. Furthermore, this result can be useful for a deduction on the past, not for a prediction on the future. A possible interpretation is in fact to infer from the final state of the wave packet its maximal amplitude (*t* = *b*) without the third term of dispersion relation (*γ* = 0)

**Table 3.**
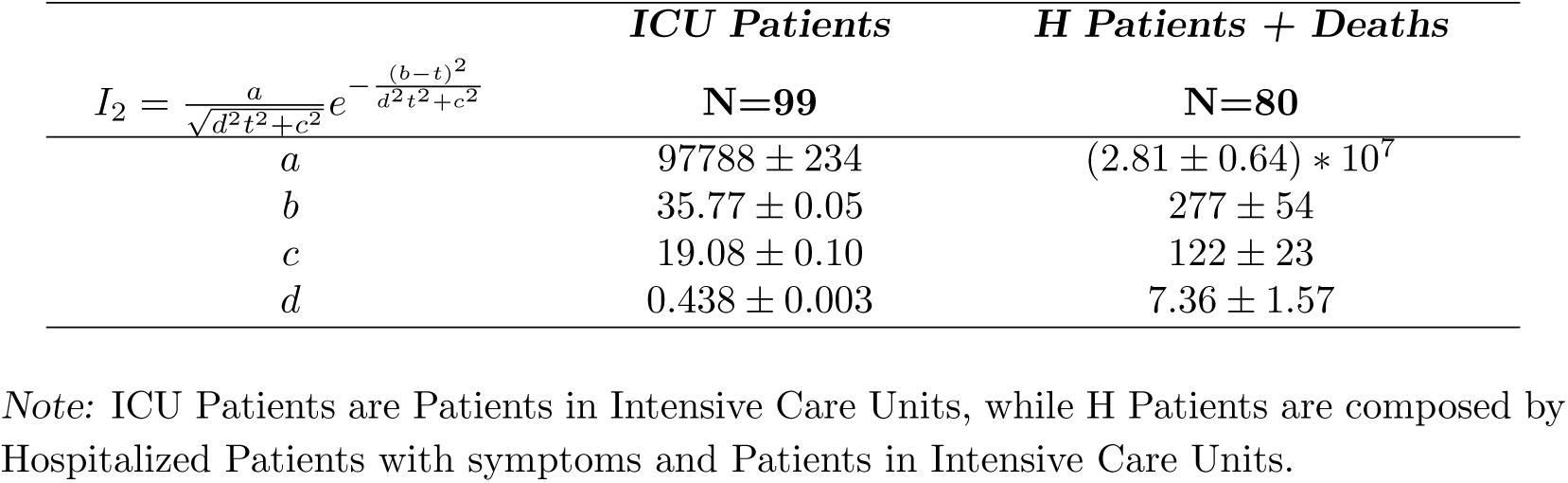
Best fit parameters for the curves in Fig. 3 of ICU Patients and H Patients + Deaths of the Epidemic Spreading.

|  | *ICU Patients* | *H Patients + Deaths* |
| --- | --- | --- |
| $I_2 = \frac{a}{\sqrt{d^2 t^2 + c^2}} e^{-\frac{(b-t)^2}{d^2 t^2 + c^2}}$ | **N=99** | **N=80** |
| $a$ | $97788 \pm 234$ | $(2.81 \pm 0.64) * 10^7$ |
| $b$ | $35.77 \pm 0.05$ | $277 \pm 54$ |
| $c$ | $19.08 \pm 0.10$ | $122 \pm 23$ |
| $d$ | $0.438 \pm 0.003$ | $7.36 \pm 1.57$ |

*Note:* ICU Patients are Patients in Intensive Care Units, while H Patients are composed by Hospitalized Patients with symptoms and Patients in Intensive Care Units.

**Figure 3a.**
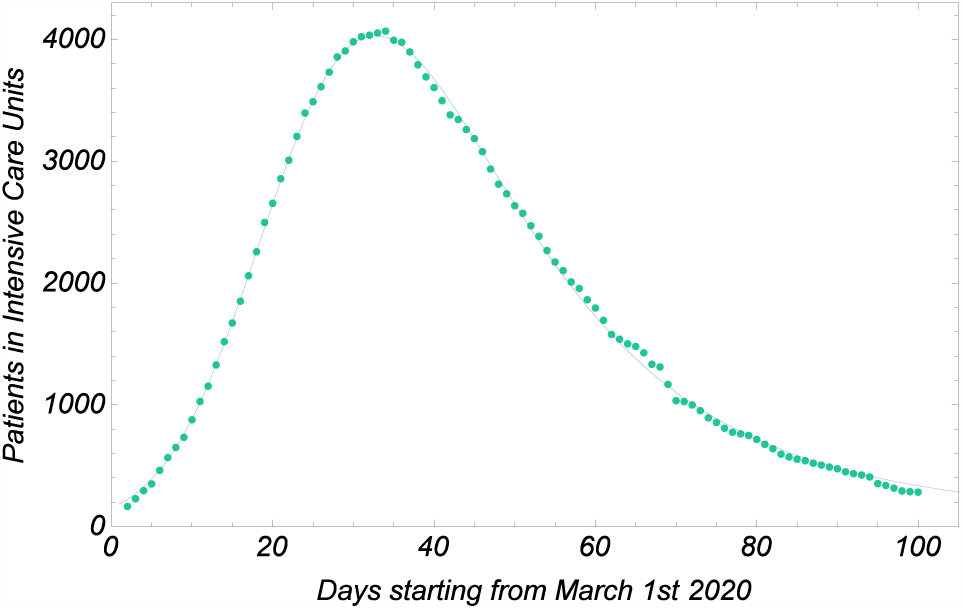
The best fit curve represents the spreading in time of a wave packet in a dispersive medium according with the function (10) in the text.

**Figure 3b.**
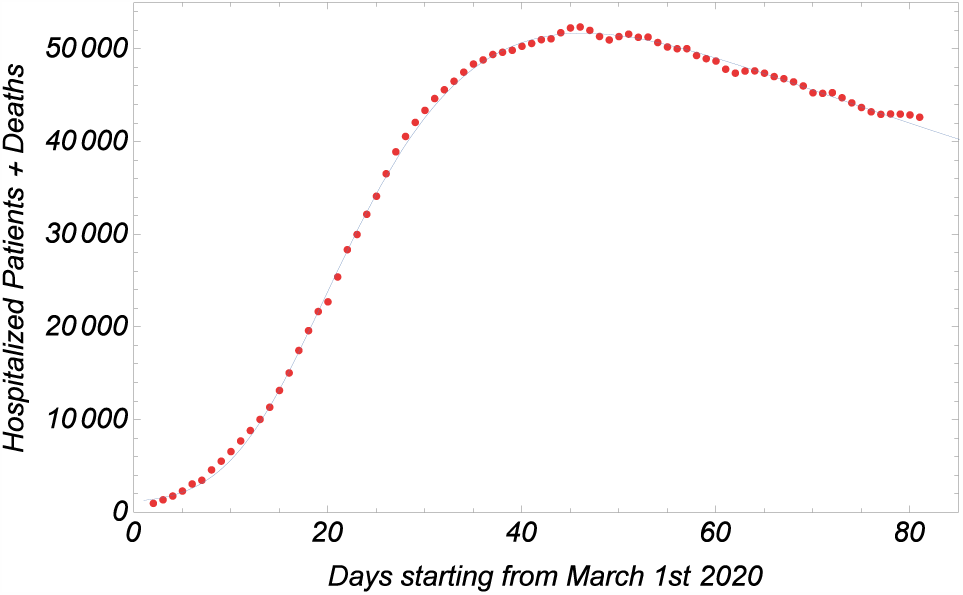
The best fit curve represents the spreading in time of a wave packet in a dispersive medium according with the function (10) in the text.

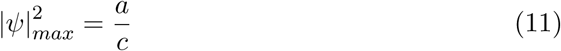

Hence, in the absence of lockdown (that is without a dispersion of the wave *Max* packet), the maximal of the first curve (**Fig. 3a**) would be 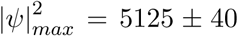 instead of the experimental value 4068 and of the second curve (**Fig. 3b**) 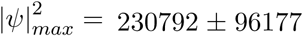 instead of 52367. If the ratio between the estimated and the real number of hospitalized plus deaths is 4.407, it is legitimate to think that, without the lockdown, the number of deaths would have been much greater than the value of 34405 on June 16th. Note also that, while the amplitude depends on *γ*, the time corresponding to the peak of the curve depends on the group velocity. If we change the medium putting *γ* = 0, also *v*_*g*_ will be different and its value will determine the date in which the curve will reach the maximal amplitude.

## 5 Conclusions

We have performed a statistical analysis of the data about the spreading of COVID-19 in Italy and we have described the trend of the number of ICU patients and of hospitalized patients plus deaths during 100 days, from March 2nd to June 9th. We have used a very simple model that describes the spreading of the virus as the propagation of a wave packet in a dispersive medium. We have found two possible functions that fit well the experimental data and can be used to predict the future behavior of the pandemic and also to infer the benefits due to the lockdown measures simulated in the model by the dispersion relation of the medium in which the wave packet propagates. The studied curves can be considered as a good thermometer of the trend of the virus in Italy and can be suggested as a useful tool to understand the evolution of the epidemic also for other countries. At this point it would be useful to understand if the physical model can work to explain the spreading of the pandemic also in other countries that have adopted similar lockdown measures.

## Data Availability

March 2nd to June 9th

https://opendatadpc.maps.arcgis.com/sharing/rest/oauth2/authorize?client_id=arcgisonline&display=default&response_type=token&state=%7B%22returnUrl%22%3A%22http%3A%2F%2Fopendatadpc.maps.arcgis.com%2Fapps%2Fopsdashboard%2Findex.html%22%2C%22useLandingPage%22%3Afalse%7D&expiration=20160&locale=it-it&redirect_uri=https%3A%2F%2Fopendatadpc.maps.arcgis.com%2Fhome%2Faccountswitcher-callback.html&force_login=false&hideCancel=true&showSignupOption=true&canHandleCrossOrgSignIn=true&signuptype=esri&redirectToUserOrgUrl=true

## 6 Acknowledgement

This work was partially supported by research funds of the University of Sannio.

## Notes

### Competing Interest Statement

The authors have declared no competing interest.

### Author Declarations

University of Sannio

## References

[1] A. Feoli, A.L. Iannella and E. Benedetto, https://doi.org/10.1101/2020.05.09.20096149 (2020).

[2] http://opendatadpc.maps.arcgis.com/apps/opsdashboard/index.html/b0c68bce2cce478eaac82fe38d4138b1

[3] D. Fanelli and F. Piazza, Chaos, Solitons & Fractals Volume 134, 109761 (2020).

[4] Z. Ceylan, Science of The Total Environment Volume 729, 138817 (2020).

[5] K. Liang, Infection, Genetics and Evolution Volume 82, 104306 (2020).

[6] X. Zang, R. Ma and L. Wang, Chaos, Solitons & Fractals Volume 135, 109829 (2020).

[7] P. Girardi, L. Greco, V. Mameli, et al., Significance, June (2020)

[8] A. Dziugys, M. Bieliunas, G. Skarbalius, et al., https://doi.org/10.1101/2020.04.28.20083428

[9] J.D. Jackson, Classical Electrodynamics (2nd edn. Wiley, New York) (2013)

[10] S. Flügge, Practical Quantum Mechanics (Springer-Verlag Berlin Heidelberg) (1999)

